# Limb state accounts for differences between motor imagery and action in motor cortex

**DOI:** 10.64898/2026.03.13.26348353

**Authors:** Samantha N. Johnson, Milan Rybář, Charles M. Greenspon, Dalton D. Moore, John E. Downey, Brian M. Dekleva, Nicholas G. Hatsopoulos

**Author notes:** co-senior authors (Brian M. Dekleva, Nicholas G. Hatsopoulos). <u>Disclosures:</u> NGH serves as a consultant for Blackrock Neurotech. CMG has received sponsored travel from Blackrock Neurotech and serves as a consultant for Neuralink Corp.

## Abstract

The motor cortex is involved not only in movement execution but also in motor imagery, a process leveraged by decoding algorithms for brain-computer interface (BCI) applications in individuals with severe motor impairments. Previous work has shown that population activity during execution and imagery occupies partially overlapping regions of neural state space while also engaging distinct subspaces unique to each motor state, suggesting that decoders trained in one condition may not generalize to the other. Moreover, movement execution likely includes neural representations of both motor output and proprioceptive feedback, which themselves may occupy distinct or overlapping regions of neural state space. To explore these distinctions, we studied two individuals with incomplete spinal-cord injuries and partial residual proximal arm function performing a center-out reaching task in three conditions: motor imagery, active execution, and passive movement. We found that decoders trained on neural activity from motor imagery failed to generalize to either active or passive movements. In contrast, decoders trained on active or passive movement activity generalized reciprocally. Population analysis revealed distinct dynamics depending on limb state and proprioceptive feedback, which could explain this lack of generalization. These results suggest that motor imagery engages motor cortical representations distinct from those recruited during actual movements, either actively or passively generated, with important implications for the design of BCI decoders.

## INTRODUCTION

Perfecting a golf swing is a complex motor skill that depends not only on physical execution but also on motor planning and sensory feedback. An experienced golfer might mentally rehearse the motion and take a few practice swings before taking a shot, whereas a novice may be passively guided through the movement by a coach, relying on sensory feedback to help them feel the correct motion. These diverse strategies of mental imagery, active movement, and passive movement illustrate the different modes through which the brain can engage with and refine motor behavior. Understanding how the brain represents and differentiates these modes of motor engagement is essential for revealing the neural mechanisms underlying motor control.

Although motor cortex is primarily associated with the execution of voluntary movement, it is also engaged during other non-execution motor activities. Neural activity related to observed or imagined movements has been observed across species, suggesting that motor cortex participates in motor intent even without executed movements ^1–6^. Motor cortical activity is also modulated during passive limb movement, largely through proprioceptive input, indicating sensitivity to changes in limb state in the absence of voluntary control ^7–12^. Prior work has further shown that these motor states can recruit overlapping neural populations in motor cortex, while also exhibiting differences in modulation patterns or temporal dynamics ^5,13^. While it has been established that motor cortex supports multiple motor control strategies, the extent of these similarities and differences across motor states remain unclear.

Recent work has sought to characterize the shared and distinct neural activity across motor states, particularly by comparing motor imagery and active movement. Using a wrist isometric force task where a human subject either imagined or actively performed the task, Dekleva et al.^13^ showed that neural population activity could be decomposed into a shared subspace and condition-unique subspaces for imagery and action. The action-unique subspace was interpreted as an output-potent dimension capturing efferent motor output as well as afferent sensory feedback. However, with no overt movement, the isometric nature of this task prevented examining the extent to which changing limb state may have contributed to these differences between imagery and action.

Here, we extend this framework by studying a proximal arm reaching task that includes passive movements in addition to imagined and active movements, allowing us to directly examine how limb state and volitional control shape motor cortical representations. Two participants with partial spinal-cord injuries who retained some residual proximal arm control engaged in a center-out reaching task while observing a virtual arm performing the same task under these three different conditions. We found that decoders trained on motor imagery were largely unable to generalize to active or passive movements, whereas decoders trained on active or passive conditions showed greater cross-condition generalization. Analyses of neural dynamics further indicated that active and passive movements share more temporal structure with each other than with imagery, consistent with the presence of limb state representations in motor cortex. When separating neural activity into shared and unique subspaces, we found that dimensions within the shared subspace were more highly correlated between active and passive movements compared to action and imagery, providing another potential explanation for the lack of cross-condition generalization involving imagery. Together, these findings highlight how motor imagery engages motor cortical representations distinct from those recruited during native limb movements.

## RESULTS

We asked two participants (C1 and C2) to observe a virtual arm performing a center-out-and-back reaching task on a television screen. Trials were performed under one of three verbally instructed conditions: imagined, active, or passive (Fig. 1a). In the imagined condition, participants imagined producing the observed movement without overt limb motion. In the active condition, participants used their native limb to replicate the timing and direction of the virtual arm’s movement. In the passive condition, the participant’s arm was passively moved to match the virtual arm while participants imagined producing the movement without intentionally assisting it. All conditions involved identical visual input and required task engagement, while differing in control strategy and somatosensory feedback: no limb movement in the imagined condition, proprioceptive input without voluntary control in the passive condition, and both proprioception and internally generated control in the active condition.

**Fig. 1.**
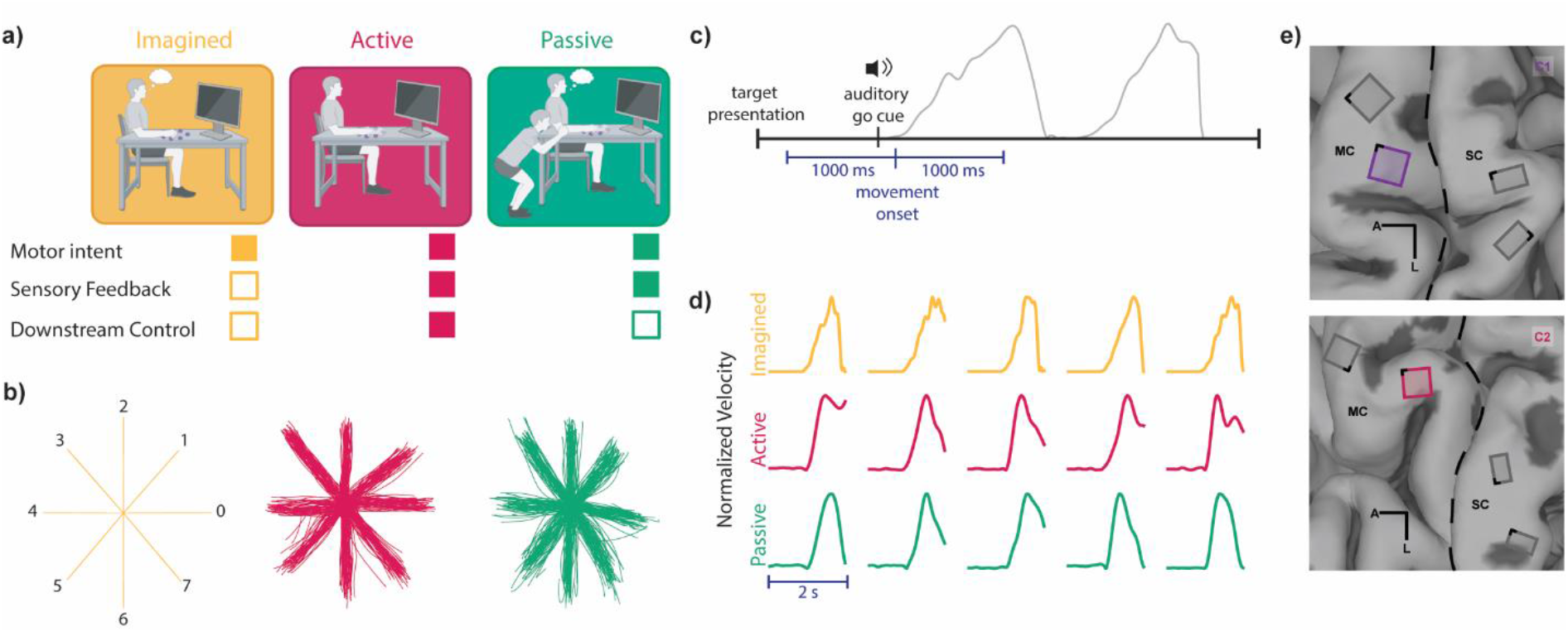
Experimental Paradigm. **a**, Participants placed their arm on a rolling sled at the center of a set of four or eight targets. They performed the center-out reach task shown by a virtual arm on the screen in front of them in each of three conditions, with an experimenter moving their arm for them in the passive task. Each of the three conditions had a unique combination of motor intent, sensory feedback, and downstream control. **b**, Position traces of the arm throughout the task, using virtual arm kinematics in the imagined condition and tracked wrist kinematics in the conditions with native limb movement. Numbers denote the target direction label. **c**, Trial structure with an example velocity trace from one trial to demonstrate timing. The blue segment designates the time window used for subsequent analyses. **d**, Velocity traces of five trials in each condition, using the 2 s window around movement onset, as shown in **c**. Noise in the imagined condition is a consequence of the MuJoCo physics engine controller and is not observable to participants. **e**, Locations of microelectrode arrays implanted in sensorimotor cortices. Only recordings from the highlighted array are used in analyses. MC and SC denote motor cortex and somatosensory cortex.

Target directions were randomized within each session (eight targets for C1; four for C2, reflecting individual movement capabilities). In the imagined condition, kinematics were defined by the virtual arm trajectory (Fig. 1b), whereas in the active and passive conditions, arm kinematics were tracked using DeepLabCut markerless motion capture (Mathis et al., 2018). Each trial began with a visual target cue, followed one second later by an auditory “go” cue, after which the virtual arm executed a three-second outward-and-return movement (Fig. 1c). Due to kinematic variability from motor impairments, analyses were restricted to a window from 1000 ms before to 1000 ms after movement onset, capturing the more consistent center-out phase of the movement (Fig. 1c,d). Each participant completed two sessions (one longer and one shorter session, with approximately 400 and 120 trials per condition, respectively). Trials exhibiting large kinematic deviations from the session-mean trajectory were excluded.

Throughout the experimental sessions, we recorded neural activity from motor cortex. Participants were implanted with four chronic NeuroPort Electrode arrays (Blackrock Neurotech., Salt Lake City, UT) spanning sensorimotor cortex as part of an ongoing clinical trial (NCT01894802) (Fig. 1e). Analyses were restricted to neural activity from the lateral motor array in the precentral gyrus in both participants.

### Task Condition Shapes Single-Channel Responses

We first asked whether single-channel motor cortical responses differed in modulation and tuning properties across imagined, active, and passive movement conditions. A modulation index was computed for each channel and condition, calculated as modulation depth across trials, and compared to baseline activity measured during a 60 s window without movement (see ‘Modulation’ in methods). Modulation increased relative to baseline in all conditions, indicating motor cortex engagement in all three motor states (Fig. 2a). However, several channels were differentially modulated depending on the condition, with some channels only being modulated in a subset of conditions (Fig. 2b, left), while others were modulated in all conditions (Fig. 2b, right).

**Fig. 2.**
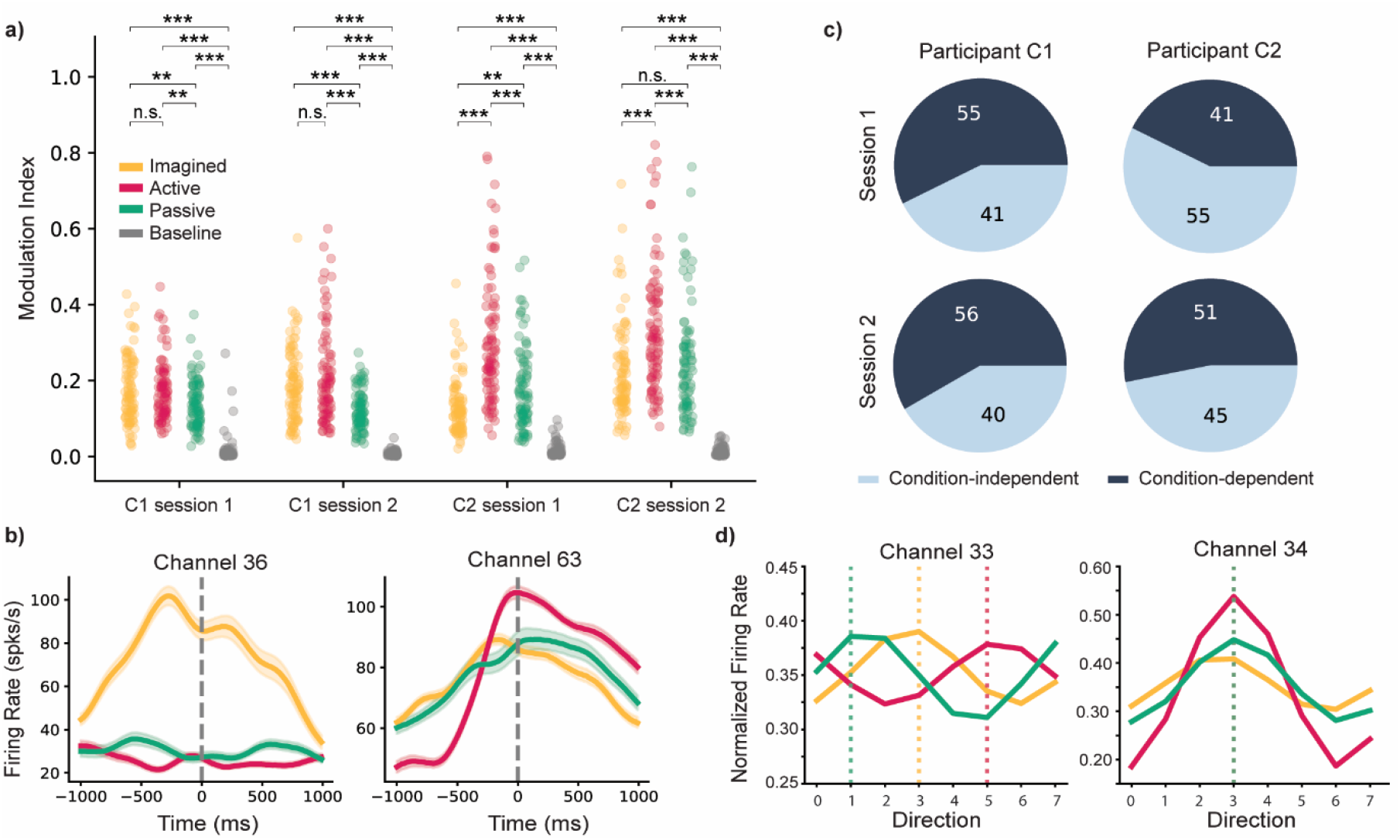
Single channel activity differs across conditions. **a**, Modulation of channel activity in each condition compared to baseline (*p<0.05 **p<0.01 ***p<0.001, Mann-Whitney U). **b**, Two example peri-event time histograms for one reach direction. The shaded regions show standard deviation across trials, and the dashed gray lines represent movement onset. **c**, Proportion of condition-dependent and condition-independent channel responses. **d**, Cosine tuning curves for two example condition-independent channels. Dashed lines show preferred direction in each condition.

Directional tuning was then quantified across channels in each condition. Channels tuned in all conditions were identified as “condition-independent,” whereas channels directionally tuned in only a subset of the conditions were classified as “condition-dependent” (Fig. 2c; see ‘Preferred Direction and Tuning Curves’ in methods). Both participants showed a mixture of condition-dependent and condition-independent channels, suggesting that partially overlapping neural populations contribute to movement representation depending on condition.

Within condition-independent channels, directional tuning curves did not necessarily match across conditions. When looking at directional tuning across conditions, we found that while some channels retained similar preferred directions in all three conditions (Fig. 2d, right), others showed different preferred directions depending on task condition (Fig. 2d, left). This indicates that even within a set of channels exhibiting tuning across conditions, the representation of movement direction is context sensitive, with tuning properties flexibly adapting to behavioral demands.

### Lack of generalization between imagined and native limb movements

After establishing modulation and tuning differences between single channels in various motor states, we next looked to determine if these differences between conditions could be observed at the population level. When decoding from neural activity on each trial using multinomial logistic regression to classify the task condition (imagined, active, or passive), all three conditions could be reliably distinguished (success rate = 0.98), demonstrating that neural representations underlying each condition are distinct and identifiable. Together with the presence of partially overlapping responses of neural populations across contexts, this suggests that condition-dependent differences could limit decoder generalization.

To evaluate cross-condition generalization, three condition-specific Kalman filter decoders were trained to predict instantaneous velocity throughout the trial and tested across all three conditions (see ‘Velocity Regression Decoding’ in methods). Performance was highest when decoders were evaluated on their training condition (Fig. 3a). Notably, cross-condition generalization between active or passive conditions performed well, indicating similar neural structure between these overt limb movement conditions. In contrast, the imagined condition showed significantly less cross-condition generalization to conditions involving native limb movement (C1: p < 0.001, C2: p<0.001, Mann-Whitney U test). Decoding failures in all conditions manifested in multiple ways, including temporal offsets, mismatched amplitude or speed, and even incorrect directionality (Fig. 3b).

**Fig. 3.**
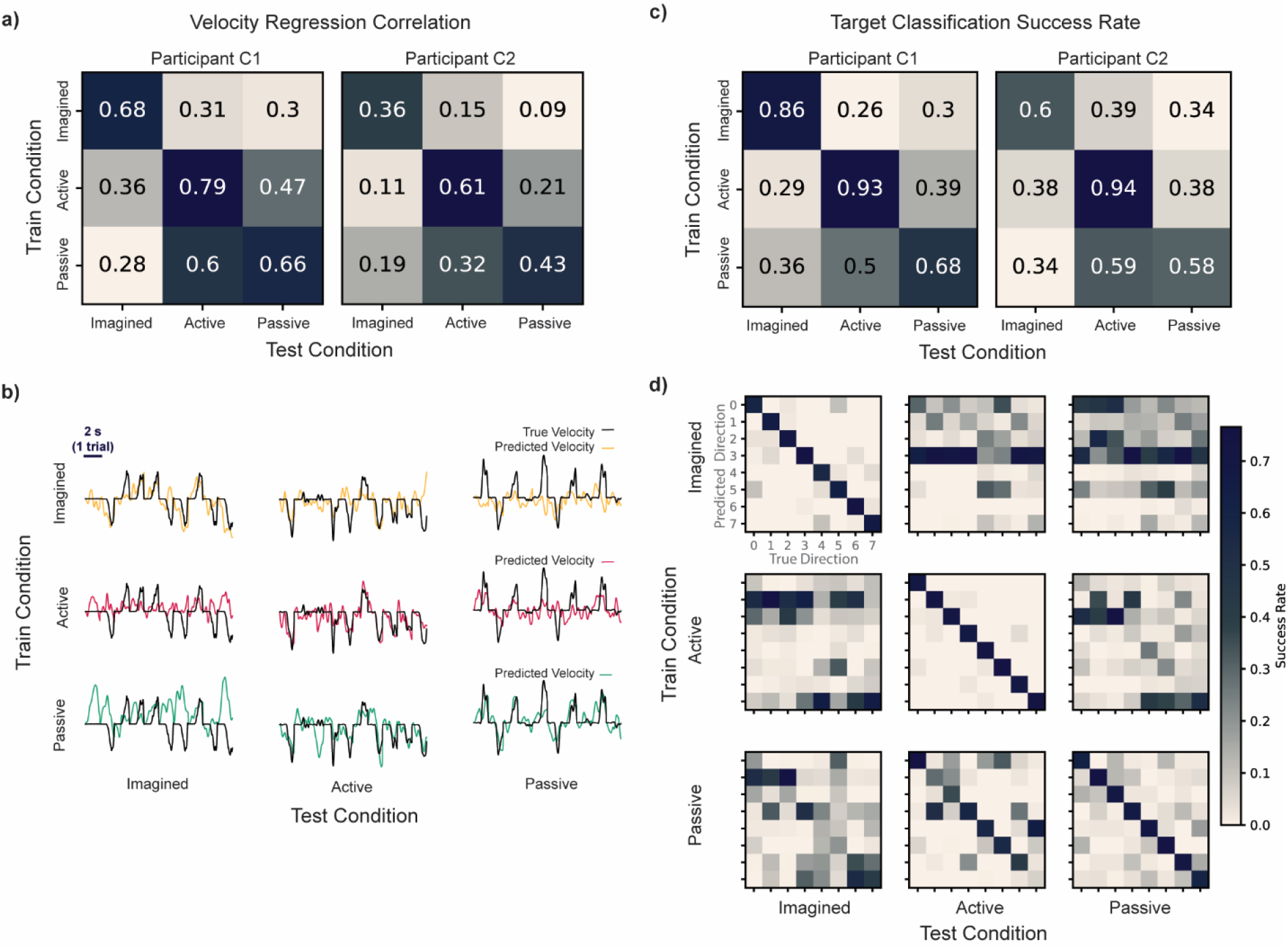
Wiener Filter Classification and Kalman Filter Regression Decoding. **a**, Velocity regression generalization across conditions for both participants. Rows show decoders trained on each of the three conditions, and columns show performance when tested on each condition, with the off-diagonal showing cross-condition generalization. Performance is reported as Pearson’s r correlation, averaged across x and y dimensions. **b**, Velocity prediction traces from participant C1 of the x-dimension from 10 concatenated trials. Black trace shows true velocity, while colored trace shows predicted velocity, with color corresponding to training data for each decoder. Most trials have a change in true velocity, but some trials have a constant true velocity, as this is a 2-dimensional task and therefore cardinal directions up/down will have no change in x velocity. **c**, Same as **a**, but target location classification by Wiener filter, reported as classification success rate. **d**, Confusion matrices for classifier performance of participant C1, with each trained decoder as a row and test conditions as columns in the larger 3×3 array. Target directions match the labeled positions in **Fig. 1b.**

Decoding instantaneous velocity in two dimensions requires precise prediction of a continuous, noisy signal, which can make generalization across conditions challenging. The failure of velocity decoders to generalize in this setting suggested that neural representations of fine-grained kinematics differ substantially between imagery and native limb movements.

However, it remained possible that more abstract, high-level task variables, such as target direction, were preserved across conditions even when kinematic representations were not. To assess this possibility, three separate decoders using Wiener filter classification were trained to decode target identity on individual trials (see ‘Target Classification’ in methods). As in the velocity regression, the imagined classifier could not generalize to conditions with native limb movement, while active and passive classifiers could generalize across native limb conditions (Fig. 3c; C1: p < 0.001, C2: p<0.001, Mann-Whitney U test), although passive generalization to active is notably better than the reverse. Examining the confusion matrices, classifiers trained on the imagined condition show more consistent failures, where the decoder struggles to find distinctions between directions and often predicts the same target (Fig. 3d). Together, these decoding results indicate that limited generalization between imagined and native limb movement conditions is not explained by decoder task complexity but rather reflects differences in the neural representations of both kinematics and abstract task features.

### Dynamic population structure distinguishes native limb movement from motor imagery

Decoding of both velocity and target identity revealed a fundamental difference in neural representation between imagined movements and physical movements, whether actively or passively performed. To directly examine these representational differences, we compared the dynamics of the neural population throughout the course of the trial. We trained a Wiener filter on each condition to decode target identity using only neural activity during peak velocity and tested decoding performance in other temporal windows of the trial (Fig. 4a; see ‘Sliding Window Temporal Classification’ in methods). This revealed that the stability of task representations throughout the trial varies between conditions. In the imagined condition, the decoder trained on the peak velocity window could perform well across the entire trial, indicating consistent neural representations in all phases of the task. However, in both the active and passive conditions, the decoder performed worse earlier in the trial, particularly before movement had started. As this performance corresponds to the change in limb state over time, we reasoned that the representation of limb state in physical movements may explain motor state differences, especially the pronounced differences between imagined and native limb movements.

**Fig. 4.**
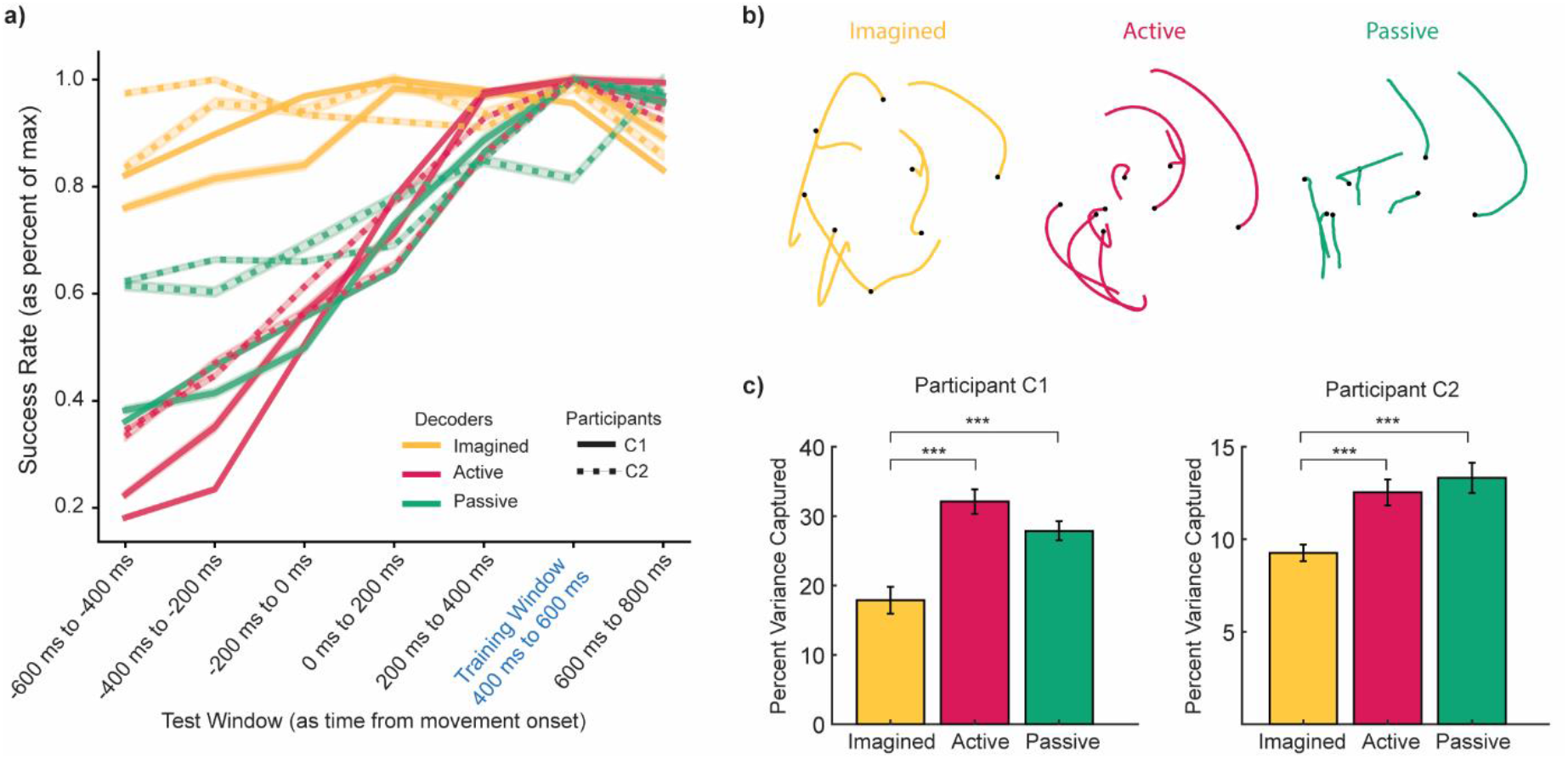
Neural dynamics reflect native limb movement. **a**, Sliding window Wiener filter classification. The blue text highlights the training window in all three conditions. The shaded region represents the standard error the mean. **b**, Neural trajectories in the first plane from the jPCA dimensionality reduction for participant C1, where the black dot represents the earliest time point in the series. **c**, Percent variance captured by the first two jPCs. Error bars show standard deviation of the jackknife cross-validation (***p<0.001, Mann-Whitney U).

The question remains, however, whether the structure of these time-varying representations changes similarly between active and passive movements. To investigate this, we examined the rotational dynamic structure of each condition, which has a well-documented presence in active reaching movements (Churchland et al., 2012; Suresh et al., 2020; see ‘Dynamical Systems Analysis’ in methods). In the active movements, we see rotational dynamics of the neural trajectories characteristic of upper limb reaches (Fig. 4b). The leading rotational dimensions explain significantly higher variance in the active and passive movements compared to the imagined movements (Fig. 4c; p < 0.001, Mann-Whitney U test). Thus, the structure of neural representations during active and passive movements can both be better explained by rotational dynamics, while that structure is not as pronounced in the absence of native limb movement. This rotational structure may contribute to the time-dependent changes in classification observed during native limb movements.

### Population activity comprises shared and unique subspaces

Previous work exploring action and imagery found that the population response could be decomposed into condition-unique and shared subspaces ^13^. The condition-unique subspaces constitute activity patterns that are active during one condition but not another, while shared subspaces are active across conditions. Applying this technique to our data, we found that all three conditions were composed of both unique and shared subspace activity (Fig. 5a; see ‘Subspace Decomposition’ in methods), indicative of a population structure that utilizes both a common substrate (shared subspace) as well as dimensions unique to each volitional state.

**Fig. 5.**
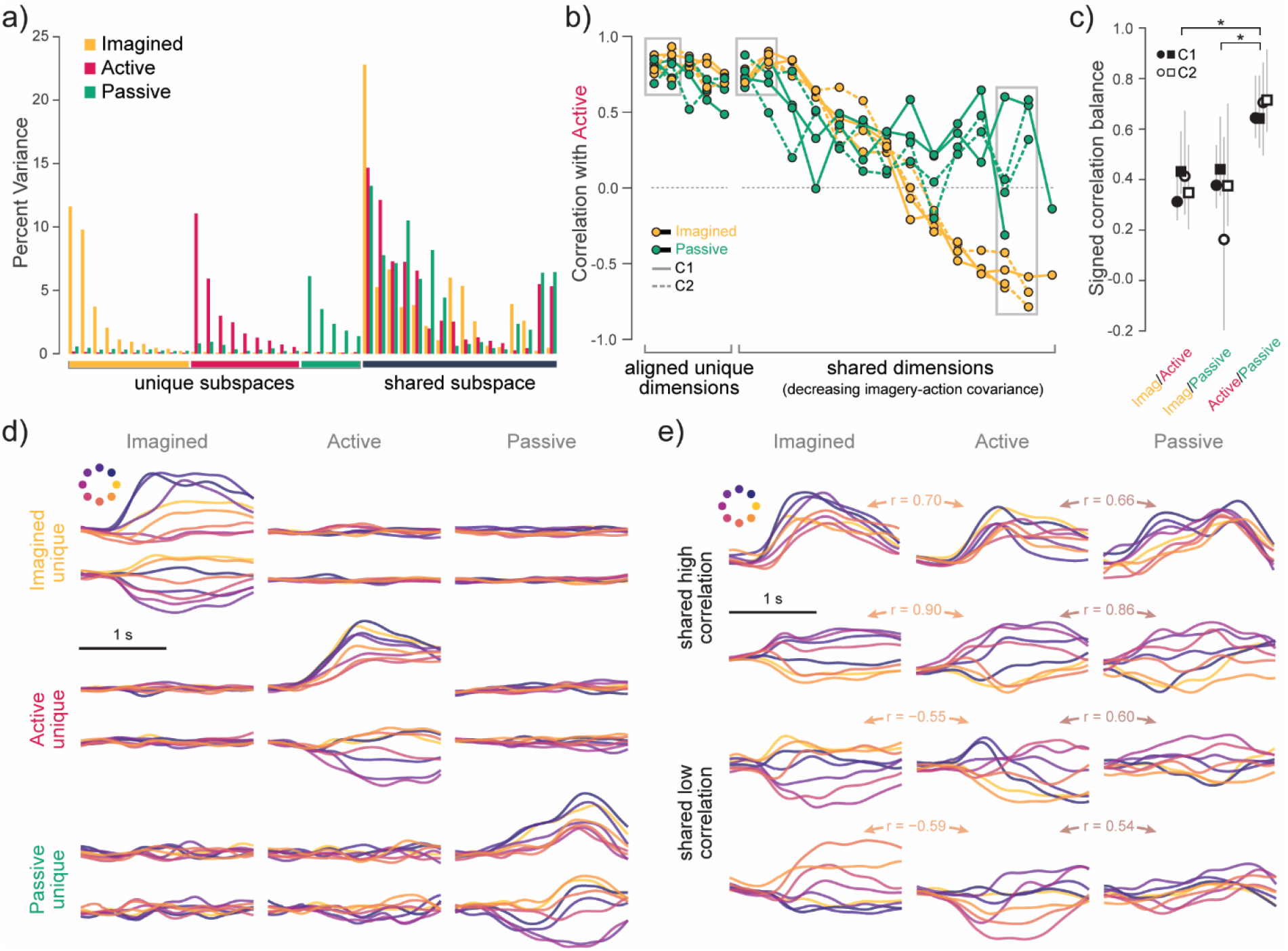
Comparison of subspace dynamics across unique and shared subspaces. **a**,Example subspace decomposition. We reduced the population activity using principal components analysis and then found an orthonormal transformation that separated subspaces with condition-specific activity (unique) from those with multi-condition activity (shared). **b**, Correlation of subspace dynamics across condition pairs (active/imagined and active/passive). Comparisons across unique subspaces were done following Procrustes alignment. Comparisons within the shared subspace were done on matched dimensions, ranked by imagery-action covariance. Gray boxes indicate examples displayed in panels **d** and **e. c**. Fraction of positive cross-condition covariance in the shared subspace. Error bars represent 95% confidence intervals following bootstrapping across trials. *p<0.05, bootstrap difference test. **d**. Example dynamics within the unique subspaces for all three conditions. The two dimensions of each subspace are indicated by the leftmost box in **b. e**. Example dynamics in the shared subspace. Top two rows reflect dimensions of the shared subspace with high correlation between both active-imagined and active-passive, as indicated by middle gray box in **b**. Bottom two rows are dimensions with high active-passive correlation but negative active-imagery correlation, as indicated by rightmost gray box in **b.**

From the decomposed subspace activity, we aimed to characterize the similarity in dynamics across conditions. We first calculated the correlations on individual dimensions of the aligned activity (Procrustes) between the active and imagined conditions, ranking the dimensions by decreasing covariance. We found that the unique subspace responses were uniformly highly positively correlated (Fig. 5b,d), indicating that each one comprised a very similar set of dynamic features during its respective volitional state. However, dimensions within the shared subspace showed more variability in dynamic similarity across conditions. We observed a broad range of response similarity between action and imagery, from strongly positively correlated to strongly negatively correlated (Fig. 5b,e). However, when we computed the correlation between active and passive conditions on those same dimensions, we found that they were predominantly positive. That is, some action-related responses within the shared subspace change significantly during pure imagery but are recovered when imagery is supplemented with passive limb movement.

To further examine the incidence of negative correlations within the shared subspace, we computed a signed correlation balance metric for each pair of conditions. This metric is defined as the squared-correlation–weighted average sign of matched dimension correlations, giving a bounded measure of whether coupling is predominantly positive or negative. We found that the signed correlation balance was significantly higher between the active and passive conditions compared to imagery/active or imagery/passive (Fig. 5c). This indicates that shared subspace activity is more consistent across volitional states when they share similar limb states or limb movement (i.e. active and passive).

### High-level task features are represented in the shared subspace

By construction, the shared subspace contains dimensions that explain meaningful variance during all conditions. In contrast, the orthogonal unique subspaces explain activity only during a select task. This raised the possibility that the shared subspace may be better able to generalize between conditions compared to the full neural activity, which would contain unique dimensions for each motor state. However, when decoding either target identity or instantaneous velocity from the shared subspace, we again find that there is poor generalization between imagined movements and native limb movements (Fig. 6a,b). When compared to decoder performance on full neural activity, despite optimizing for a shared signal, we do not see improvements in generalization when using the shared subspace (Fig. 6c; p=0.06, p=0.13, two-tailed Wilcoxon signed-rank test). Therefore, although the shared subspace contains most of the total variance across tasks, these signals are still insufficient to generalize between limb state.

**Fig. 6.**
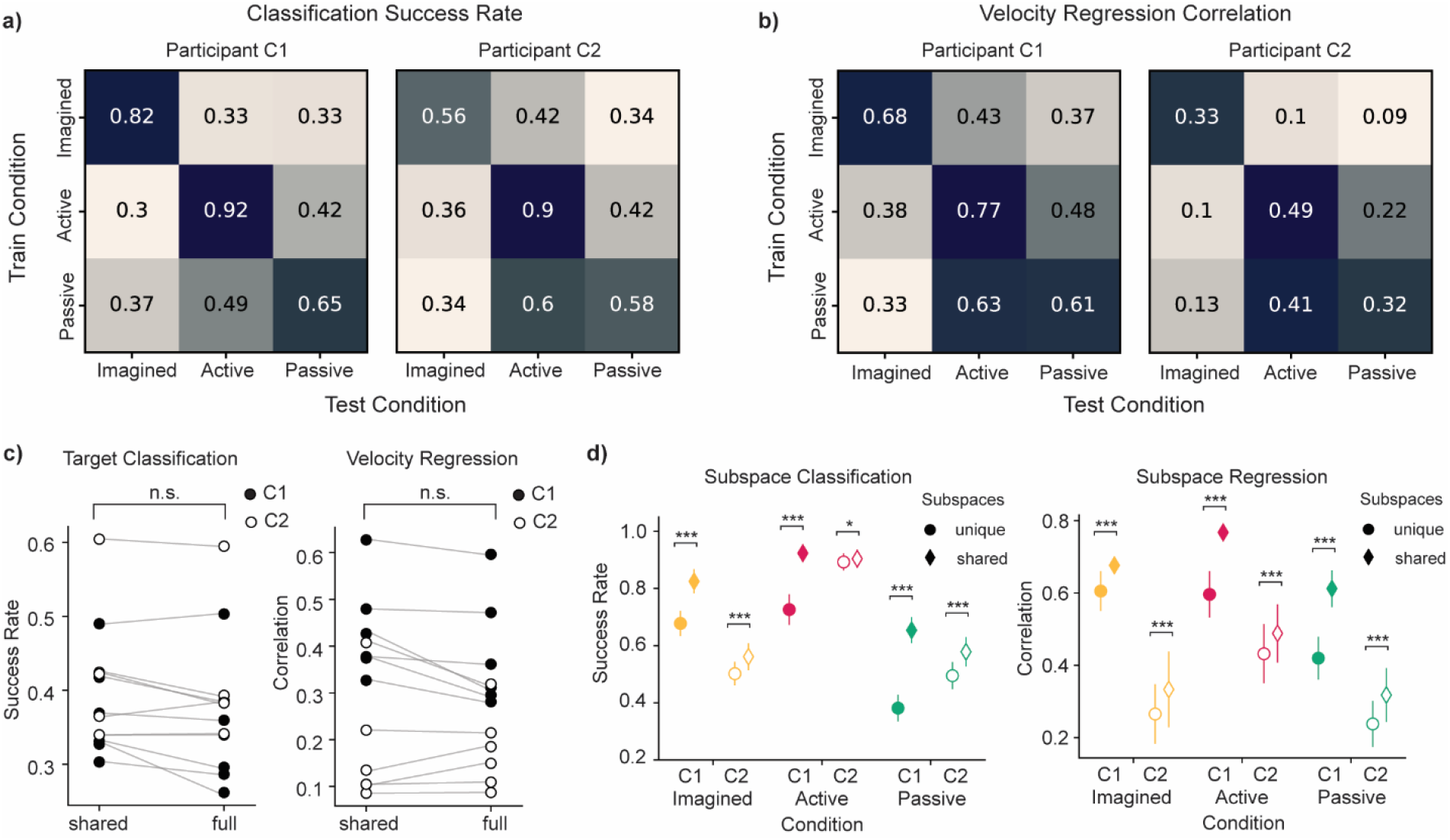
Decoding from shared and unique subspaces. **a**, Wiener filter classification success rate when decoding from the shared subspace, same format as **Fig. 3c. b**, Kalman filter velocity regression decoding from the shared subspace, reported as correlation, same format as **Fig. 3a. c**, Comparison of off-diagonal cross-condition generalization from the shared subspace **(Fig. 6a,b)** and the full neural activity **(Fig. 3a,c)** (not significant, two-tailed Wilcoxon signed-rank test). **d**, Decoding performance compared when training on unique or shared subspaces. Error bars show standard deviation across training epochs (*p<0.05 ***p<0.001, Mann-Whitney U). Wiener filter target classification is on the left, and Kalman filter velocity regression is on the right.

To determine what information may be stored in unique subspaces relative to the shared subspace, we compared decoding performance between the two subspaces on target classification and velocity regression (Fig. 6d). In all conditions, we see that the shared subspace better decodes the instantaneous velocity of the arm as well as the high-level task feature of target identity (p<0.05, p<0.001, Mann-Whitney U). We consider velocity and target identity to be abstract, high-to-mid-level task features, distinct from downstream control directly involving muscle activation. Therefore, the better performance of the shared subspace compared to the unique subspace suggests that high-level task features are represented in the shared subspace.

## DISCUSSION

We found that motor cortex representations were most similar between motor states with native limb movement. We see evidence of this through cross-condition decoding generalization, where active and passive reaches achieve good cross-condition decoder performance. While both pure motor imagery and our passive reaches contained an “imagined” engagement with the movement, the addition of proprioceptive feedback in the passive condition likely produces an added cross-condition signal improving generalization. This indicates not only the importance of feedback in motor cortex but introduces the potential of limb state representation in the population activity. We briefly examined passive movements with no imagined component (termed “passive blind”) in one participant (Supplementary Fig. 1), where we again saw poor generalization to imagery and better cross-condition generalization with active and passive combined with imagery conditions, but with overall weaker classification performance. These results suggest that proprioception alone, even without motor intent, produces representations in motor cortex that are more similar to active movement, though future work is needed to better isolate these proprioceptive contributions.

Neural representations evolved throughout the trial when the limb was in motion. In contrast to imagery where representations remained static, conditions involving native limb movement exhibited population activity that changed across the trial, consistent with a continuously evolving representation of limb state. This distinction highlights a fundamental difference between action and imagery: when the limb moves, motor cortex activity reflects not only motor intent but also the ongoing physical state of the limb itself. Such time-varying structure naturally motivated an analysis of neural dynamics and whether these evolving representations manifest low-dimensional, structured trajectories.

The prevalence of rotational dynamics in active arm reaches has been widely shown and established as a model to explain motor cortex population activity ^14–16^. However, while proprioceptive information has been shown to contribute to these rotational dynamics ^17^, it was unclear whether passive movements in isolation produce rotational dynamics independently, and how similar any such dynamics may be to active movements. We find rotational dynamics in motor imagery in humans to explain a low percentage of variance with weak rotation metrics.

However, we find that the variance explained by rotational dynamics in passive movements was similar to that in active reaches. Thus, the changing dynamics in this rotational plane capture limb state as well as motor execution intent, and that limb state information is critical for defining neural population structure during reaches. This structural difference could be a contributing factor to poor cross-condition decoding generalization.

The subspace decomposition revealed additional complex dynamics that could also explain poor cross-condition decoding. In particular, activity in the shared subspace showed predominantly positive correlations between active and passive responses, indicating more similar dynamics than those observed between imagery and active responses. When compared to our earlier work involving an isometric wrist extension task, these differences become more interpretable. In Dekleva et al., both the shared and the unique subspaces contained only highly correlated dimensions between active and imagery conditions. However, in this current paradigm, the limb state was not fixed. While correlations remained high across all unique subspaces, action and imagery had a broader range of correlations in the shared subspace. Instead, the most positive correlations were between passive and active movement—conditions that involve a changing limb state. This suggests that the high imagery-active correlation observed previously in the shared subspace may in part arise from the fixed isometric limb configuration. However, under conditions where the limb state evolves during the task, the shared subspace contains meaningful information about the limb state itself.

With the addition of a passive condition, we can aim to better understand the biological relevance of these shared and unique subspaces. In Dekleva et al., we previously hypothesized that the shared subspace may contain high-level task information. In the present center-out reach task, target direction can be well classified from the shared subspace, supporting this claim of high-level information in the shared subspace. However, we also see the best performance of velocity regression from the shared subspace, indicating that mid-level information such as velocity also seems to be included in the shared subspace along with high-level features. We also previously suggested that the unique subspaces may contain activity relevant to downstream control, as isometric wrist force could be decoded best from the action-unique subspace. Our results again support this hypothesis, interpreting the unique subspaces as crucial to low-level information that is not captured by velocity, but may be captured by force or muscle activity.

However, without any direct measurement of force or muscle control such as electromyography (EMG), we cannot confirm from this task that low-level downstream control might be represented in these unique subspaces.

We observed a number of differences between participants, particularly in modulation and decoding of the imagined condition. One possible explanation for these differences is that participant C1 has been part of this clinical study for several years longer than participant C2, resulting in overall more exposure to imagined tasks. It has been reported that human participants frequently report needing to “learn” the skill of controlling effectors through imagination alone, and performance tends to improve with practice ^18,19^. The longer exposure for C1 could have led to higher modulation and decoding performance in the imagined condition, as this skill of motor imagery improved over time. Differences in residual movement capacity and proprioception between participants could have also contributed to differences between the two participants in the passive condition (Supplementary Fig. 2).

Understanding differences between motor states is not only scientifically important but also holds practical relevance for motor learning. Consider the earlier example of perfecting a golf swing through mental imagery, passive movements, and active rehearsal. Previous work has shown that passive movements can facilitate motor learning ^20^. Our findings provide a potential neural explanation for this effect. Passive movements produce activity in motor cortex that more closely resembles active movement. As a result, passive training may engage the motor system in a manner that is more congruent with neural patterns ultimately required for active movement. In practice, this suggests that incorporating passive movement training paradigms into motor skill acquisition may support more effective learning than imagery alone.

Beyond motor learning, these insights also have implications for BCI research. The observation that motor cortex is engaged during motor imagery has become foundational for BCI research, particularly in contexts where overt movement is limited or absent and decoders must rely entirely on imagined actions. However, incorporating passive movement with BCI control, such as with exoskeletons or functional electrical stimulation (FES), can enhance performance when the kinesthetic feedback is congruent with the user’s intended movement ^21^. Our results emphasize the importance of congruent proprioceptive feedback and highlight the potential benefits of including passive movement when training BCI systems that use native limb movement, including exoskeletons or FES. Doing so could not only advance decoding performance but would make control more biomimetic by leveraging passive representations that are closer to active movements. As a result, incorporating passive movement paradigms into BCI control may provide a more biomimetic foundation for decoder design, bringing artificial BCI control closer to the neural processes that underlie natural native limb movement.

## METHODS

This study was conducted as part of a registered clinical trial (NCT01894802) for the study of intracortical BCI under an Investigational Device Exemption from the U.S. Food and Drug Administration. The studies were approved by the Institutional Review Boards at the University of Chicago and the University of Pittsburgh. Informed consent was obtained prior to conducting any study-related procedures.

### Participants

Participant C1 (m), 55-60 years old at time of implant, presented with C4-level ASIA D spinal cord injury (SCI) that occurred 35 years prior to implant. Participant C2 (m), 60-65 years old at time of implant, presented with C4-level ASIA D SCI and right brachial plexus injury that occurred 4 years prior to implant. We chronically implanted four Blackrock NeuroPort Electrode arrays (Blackrock Neurotech, Salt Lake City, UT, USA) in each participant, with two arrays in Brodmann’s area 1 of the somatosensory cortex and two arrays in the motor cortex, one medial and one lateral ^22^. The arrays in the somatosensory cortex were 2.4mm x 4mm, with 32 1.5 mm recording electrodes in a checkerboard pattern. The arrays in the motor cortex were 4mm x 4mm with 96 1.5mm recording electrodes. Signals were recorded using the NeuroPort system at 30kHz, high-pass filtered with a 1st order 750 Hz filter, and every crossing of a voltage threshold (-4.5 RMS) was counted in 20ms bins and used for offline analysis and online decoding.

### Data Collection

Participants were seated in front of a television screen and performed variations on a center-out reaching task. The visual cue stayed the same for every condition. For each trial, the visual cue showed an arm hovering over the center of a table, where a target would appear in one of the possible target locations in a circle around the arm. Each participant rested their arm on a cushioned rolling platform to help facilitate movement along a plane while eliminating sliding friction to reduce fatigue and enable more trials to be completed.

Participants performed this task in three main conditions: imagined, active, and passive. In addition to these conditions, participants completed a passive-blind condition, where they did not know the location of the target throughout passive movement and thus did not produce any motor imagery (Supplementary Fig. 1). We found these two passive conditions to be relatively comparable and chose to focus on the passive imagined for narrative continuity.

Position of the native limb was found using DeepLabCut ^23^ to track the wrist on two overhead cameras (Blackfly S, Teledyne FLIR), down sampled to 50 frames per second. Because arms were resting on a platform with closed or relaxed fists, there was no consideration of wrist angles, aperture, or grasp in this task. Triangulation of the two cameras was completed using Anipose ^24^ to get position in 3D space. Trials with significant obstruction or poor prediction were excluded from data.

### Preprocessing

For all imagined, active, and passive movements, trials were aligned at the start of movement. For active and passive conditions, this was calculated as 10% of the peak velocity from the positions of the native limb. For the motor imagery, choosing the movement onset proved to be more difficult, as there was no native limb movement onset. We considered using the movement onset of the virtual limb as a proxy for movement onset. However, we saw two key pieces of evidence against that alignment. First, in the active condition, we see that the timing of the virtual arm and native limb have a significant offset, with the native limb often leading the virtual arm by an average of 200-400ms. Additionally, neural trajectory separation by target condition happened substantially before kinematic arm movement onset as well.

Therefore, in the absence of this ground truth, we decided to use neural movement onset as the best approximation for “movement” onset in motor imagery. Kinematics were lowpass filtered with a 4^th^-order Butterworth filter at 6Hz. Velocity was normalized between -1 and 1 for all conditions to eliminate scaling as a potential confound for generalization.

Due to concerns about biologically unfeasible neural activity from widespread noise on the medial motor array, only neural activity collected from the lateral motor array was used in analysis for both participants C1 and C2. All neural activity was filtered using a Gaussian filter with 100ms standard deviation and binned in 20ms time bins. The window of 1000ms before movement onset to 1000ms after movement onset was used for most analyses. This window was chosen to include the preparation and most of the center out movement without any of the center-back component of the reach.

### Modulation

Modulation of individual channels was calculated in each condition using modulation depth *M*_*c*_, defined as:

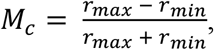

where *r* is the trial-averaged firing rate of channel *c*. For baseline modulation, we used neural activity during a 60 s no-movement window, broken up into trials of 2 s to match task trial structure. These 2 s windows were averaged, and channel modulation was calculated using the modulation depth metric defined above. Significance between condition distributions was calculated using a Mann-Whitney U test.

### Preferred Direction and Tuning Curves

Given that one participant had only four unevenly spaced target locations, cosine tuning curves were not appropriate for determining tuning. To determine if channels were context-dependent or context-independent, we instead looked for significantly different response properties across conditions. For each channel, we calculated the distribution of maximum firing rates on all trials of each direction independently. A Kruskal-Wallis test was then used to check if activity was significantly different between directions, with a criterion of *ϵ*^2^≥0.1 for medium-large effect sizes. Channels that met the effect size threshold in all conditions or no conditions were considered “Condition-Independent”, while channels that had large effect size in only one or two conditions were considered “Condition-Dependent” channels.

Preferred directions of context-independent channels in the 8-direction paradigm were calculated using cosine tuning curves fit to the average firing rate of a channel at each direction between -500 ms to 500 ms around movement onset. Cosine-fit average firing rate was normalized across all channels for visualization. For Fig. 2d, only the 8-direction paradigm was used to have better precision in preferred direction.

### Velocity Regression Decoding

To predict kinematics, we applied the Kalman filter, using implementation published by the Kording lab ^25^. The Kalman filter is a commonly used filter for kinematic decoding ^15,26–28^, as it describes kinematic dynamics using a linear relationship between past and future states. This filter used input neural data across all channels at each time point and produced kinematics at every time point in the trial, using all default parameters. Output kinematics were two-dimensional velocity, which were often averaged together for visualization, but decoded separately. We again trained three models, one for each condition. Multiple values for lag were tested but reported values used no lag due to limited performance gains and different optimal lags across conditions. Models were validated using bootstrapping with 100 resamples, where 80% of trials were used for training data and the remaining 20% of trials were used to test the model. Performance of the model was calculated as correlation between actual and predicted x and y velocities. Statistical significance was calculated using Mann-Whitney U test between two distributions, one of all four imagery cross-condition generalization scores from each of the 100 resampled bootstraps, and a second distribution made up of both active/passive cross-condition generalization scores from each of the 100 resampled bootstraps.

### Target Classification

We implemented a Wiener filter to classify the target identity in each trial from the neural activity. Depending on the participant, the output was either one of eight (C1) or one of four (C2) targets depending on the participant, with chance performance of 12.5% or 25%, respectively.

Three different models were trained, one on neural activity from each condition, implemented using the Neural Decoding package published by Glaser et al., 2020. and default parameters, which leverages scikit-learn logistic regression. Input data was an array of all channels by time points from -1000 ms to 1000 ms for each trial. Models were validated using bootstrapping with 100 resamples, where 80% of trials were used for training data and the remaining 20% of trials were used to test the model. Performance of the model was calculated as success rate, with all incorrect classifications being equally penalized, regardless of positional relation to the correct target label. Statistical significance was calculated the same way as in the Kalman Filter Regression.

### Sliding Window Temporal Classification

To determine how the neural representation changed throughout a trial, we again used a Wiener filter for classification of target location. Using 200 ms windows, we chose a single training window in each condition to be the 400 ms to 600 ms window after movement onset, which most often contained the peak velocity in a trial. That decoder, one for each condition, was then tested in other 200 ms windows spanning -600 ms to 800 ms around movement onset. Success rate was reported as percent of maximum success rate for better visual comparison across conditions. Without normalization, the maximum success rate for each condition (imagined, active, passive) of each day was as follows:

**Supplementary Table 1:** Maximum success rate of Temporal Classification before normalization.

|  | **Imagined** | **Active** | **Passive** |
| --- | --- | --- | --- |
| **C1 Session 1** | 0.717 | 0.870 | 0.593 |
| **C1 Session 2** | 0.730 | 0.797 | 0.618 |
| **C2 Session 1** | 0.404 | 0.923 | 0.489 |
| **C2 Session 2** | 0.510 | 0.908 | 0.544 |

### Dynamical Systems Analysis

Rotational Dynamics analysis was performed using the dimensionality reduction method jPCA ^14^, which finds orthonormal basis projections that capture rotational structure in neural data. We followed the preprocessing steps described in Churchland et al., 2012, including removing the cross-condition and cross-direction mean, and smoothing data using a gaussian filter with standard deviation of 60 ms. We also chose to downsample to trials with most similar kinematics based on velocity profiles. Analyses focused on the window of -60 ms to 440 ms around movement onset, which was primarily the ballistic movement of the reach.

Population data was reduced to 6 dimensions using PCA, then used jPCA to find informative planes for rotational dynamical structure. Reported variance explained and trajectory plots come from the first plane of the jPCA summary, using the first two jPCs. To estimate uncertainty in variance explained, we performed leave-one-trial-out jackknife resampling across trials. Fig. 4c reports variance explained using all trials, with error bars denoting the standard deviation across jackknife iterations. Significance was calculated using a Mann-Whitney U test between jackknife distributions between conditions.

### Subspace Decomposition

We aimed to decompose the activity collected across the three volitional states to identify condition-unique and shared subspaces. Unique subspaces represent dimensions of the population activity that are only active during a single condition, while the shared subspace contains dimensions active across multiple conditions. To do this we used a method we previously developed ^13^ called Dynamic Subspace Overlap (DySO). Briefly, we first identified each set of unique subspace responses, defined as the activity within the null space of all other conditions. For example, action-unique responses were those present within the null space of the imagery and passive conditions. Imagery-unique responses were in the null space of action and passive, etc. Once we obtained the set of all unique responses, we identified a single orthogonal subspace that recapitulated those responses. The resulting subspace was in effect the concatenation of the three unique subspaces, so we split accordingly into action-unique, imagery-unique, and passive-unique. The shared subspace was defined simply as the null space of the combined unique subspaces. In the end, the decomposition retains all geometric characteristics of the population activity (the combination of all subspaces forms an orthonormal basis set) and represents just a simple rotation that emphasizes condition-unique variance properties.

After the subspace decomposition, we performed one additional transformation to enable comparison of responses across unique subspaces. Because each condition’s unique subspace is estimated independently, their bases are arbitrary — the first dimension of the action-unique subspace has no guaranteed relationship to the first dimension of the imagery-unique subspace. Direct comparison is therefore not meaningful without first aligning them. We addressed this by applying Procrustes alignment (rotation only) to align the imagery-unique and passive-unique subspaces to the action-unique subspace, making the internal structure of each subspace as similar as possible and allowing us to compare activity patterns across conditions. This step was not necessary for the shared subspace, where each dimension is by definition consistent across conditions and can be compared directly.

### Decoding from Subspaces

Neural activity was projected into the loadings for each subspace, either unique or shared (including dimensions shared across all three conditions as well as those dimensions shared in only two conditions). Projections were mean-subtracted within condition. Using these projections as input, Wiener Filter Classification and Kalman Filter regression were implemented as described previously, with 100 epochs of 80% training data and 20% test data. A two-tailed Wilcoxon signed-rank test was used to compare cross-condition generalization between shared subspace decoding and full neural activity decoding in Fig. 6c. and Mann-Whitney U was used to calculate statistical significance in Fig 6d.

## Data and code availability

All data is stored at the Data Archive BRAIN Initiative (https://dabi.loni.usc.edu/dsi/XXXX) and code for analysis is available on GitHub (https://github.com/CorticalBionics/LimbState).

## Acknowledgements

We thank the participants for their generous contribution to the advancement of science. We would also like to thank all current and former members of the Cortical Bionics Research Group at the Universities of Chicago, Pittsburgh, and Northwestern for their contributions to this work, especially Ariana Tortolani and Lamine Camara for their early contributions. The work at the University of Chicago and at the University of Pittsburgh was supported by the National Institute of Neurological Disorders and Stroke of the National Institutes of Health under award numbers UH3 NS107714 (to NGH and JED) and R35 NS122333 (to NGH). This material is based upon work supported by the National Science Foundation Graduate Research Fellowship under Grant No. 2140001 (to SNJ).

---

**Supplementary Fig. 1.**
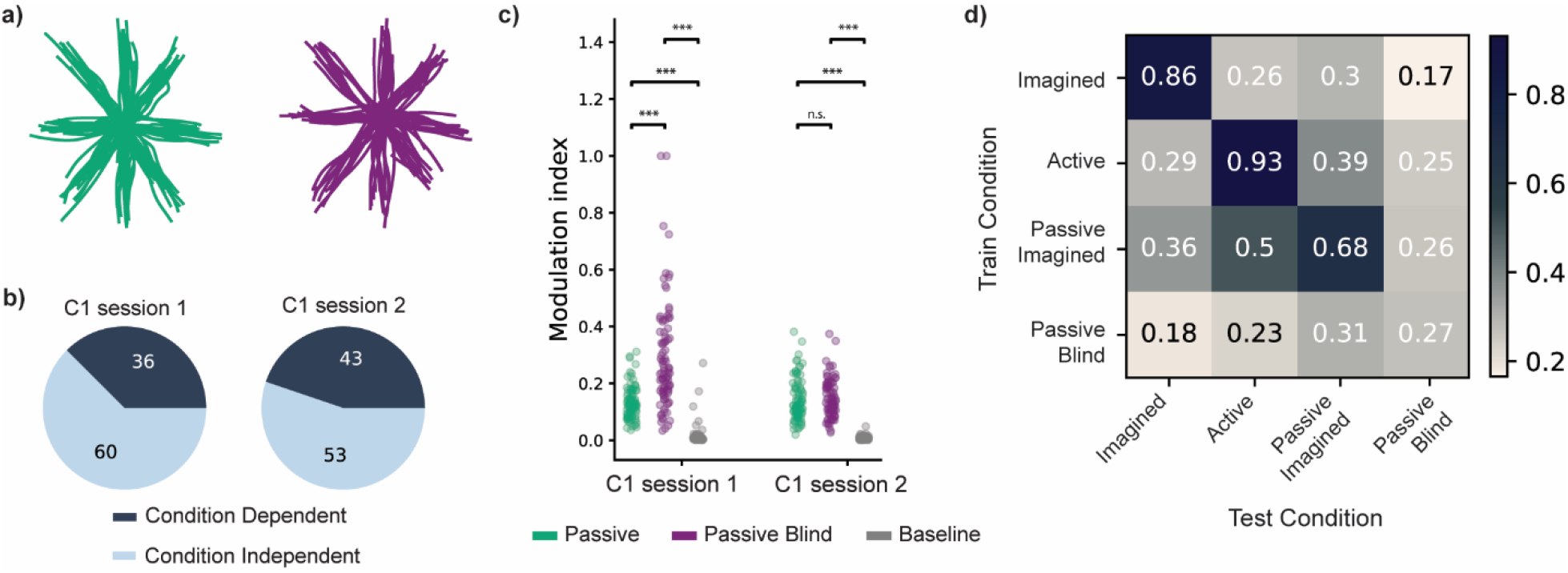
Passive versus Passive Blind condition comparison. **a**, Position traces of the arm throughout the task, using tracked wrist kinematics in both the passive + imagined (left) and passive blind (right) conditions. **b**, Proportion of condition dependent and independent channel responses. As in **Fig. 2c**, we see overlapping populations of channel responses, some that are selectively tuned to either passive or passive blind, and some that are tuned in both conditions. **c**, Modulation of channel activity in each condition compared to baseline (*p<0.05 **p<0.01 ***p<0.001, Mann-Whitney U). While significance between passive conditions is inconsistent across sessions, both passive conditions show significant modulation relative to baseline **d**, Target direction classification generalization across conditions for both participants. Rows show decoders trained on each of the three conditions, and columns show performance when tested on each condition, with the off-diagonal showing cross-condition generalization. Performance is reported as success rate. While passive blind performance is low, we see consistent trends with the passive + imagined condition, with best cross-condition generalization to the other conditions with native limb movement.

**Supplementary Fig. 2.**
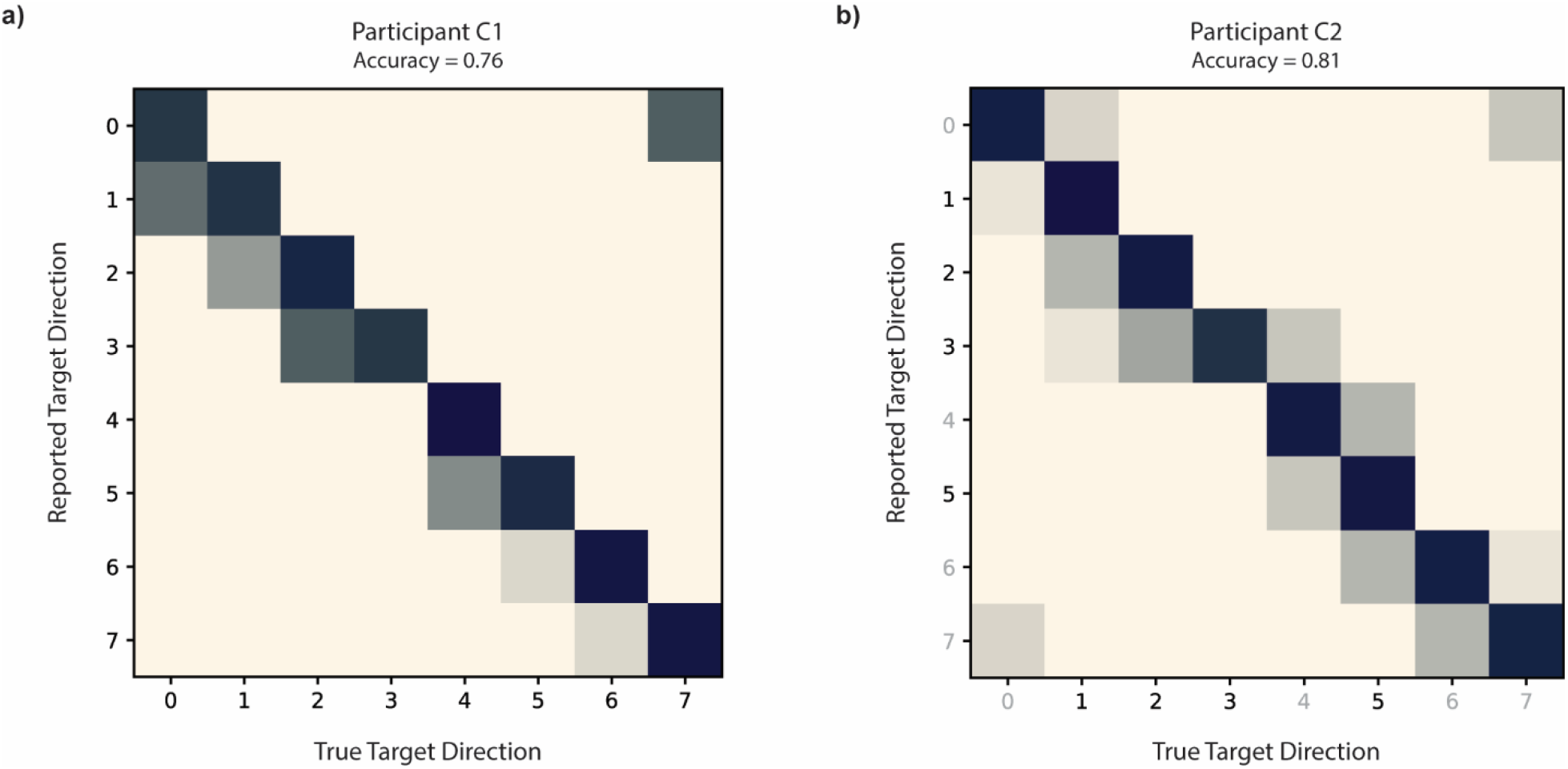
Proprioception as measured through subject reported passive classification. **a**, Confusion matrix of subject reported target direction during passive blind eight target center-out reaching task for participant C1. Participants were asked to close their eyes while their arm was moved in the same manner as the passive conditions reported previously. On each trial, the participant verbally reported target direction of their arm position. This task provided a relevant baseline for residual proprioception in each participant. Target direction labels are consistent with reported directions in **Fig. 1b. b**, Same as **a**, but for participant C2. All eight targets were tested in this experiment, while only target directions 1, 2, 3, and 5 (darker labels) were used in the main conditions due to active movement limitations.

